# Serologic evidence for early SARS-CoV-2 circulation in Lima, Peru, 2020

**DOI:** 10.1101/2024.02.13.24301472

**Authors:** Andres Moreira-Soto, Maria Paquita García, Gloria Arotinco-Garayar, Dana Figueroa-Romero, Nancy Merino-Sarmiento, Adolfo Marcelo-Ñique, Edward Málaga-Trillo, César Cabezas Sanchez, Jan Felix Drexler

**Affiliations:** Charité-Universitätsmedizin Berlin, corporate member of Freie Universität Berlin, Humboldt-Universität zu Berlin, Institute of Virology, Berlin, Germany; Tropical Disease Research Program, School of Veterinary Medicine, Universidad Nacional, Costa Rica, Costa Rica; National Institute of Health–INS (Instituto Nacional de Salud–INS), Lima, Peru; Universidad Peruana Cayetano Heredia, Facultad de Ciencias y Filosofía, Laboratorios de Investigación y Desarrollo, Lima, Perú; German Centre for Infection Research (DZIF), associated partner Charité-Universitätsmedizin Berlin, Berlin, Germany

**Author notes:** corresponding author Correspondence: Professor Dr. Jan Felix Drexler, Institute of Virology, Charitéplatz 1, 10117 Berlin, Germany, /Fax. These first authors contributed equally (Andres Moreira-Soto, Maria Paquita García).

**Keywords:** SARS-CoV-2, COVID-19, Peru, pandemic, Latin America, serology, antibodies

## Abstract

During early 2021, Peru had the highest COVID-19-associated per-capita mortality rate globally. Socioeconomic inequality and insufficiently prepared healthcare and surveillance systems likely contributed to high mortality, potentially coupled with early SARS-CoV-2 introduction. We tested 1,441 individuals with fever sampled during August 2019-May 2021 in Lima, Peru, for SARS-CoV-2-specific antibodies. Serologic testing included a chemiluminescence immunoassay and confirmatory surrogate virus neutralization testing. Early positive samples (n=24) from January-March 2020 were further tested using a plaque-reduction neutralization and avidity tests based on SARS-CoV-2 spike and nucleoprotein antigens. None of the early samples were PRNT-confirmed, in contrast to 81.8% (18/22) of a subsample from April 2020 onwards (Fischer-exact test, p<0.0001). SARS-CoV-2 antibody detection rate was 0.9% in mid-April 2020 (1/104; 95% confidence interval (CI), 0.1-5.8%), suggesting onset of viral circulation in early-mid March 2020, consistent with the first molecular detection of SARS-CoV-2 in Peru on March 6th. Mean avidity increase of 62-77% to 81-94% from all PRNT-confirmed samples during early 2020, were consistent with onset of SARS-CoV-2 circulation during late February/March 2020. Early circulation of SARS-CoV-2 was confirmed in a Susceptible, Exposed, Infected and Recovered mathematical model that projected an effective reproduction number >1, during February-March 2020. Robust serologic testing thus confirmed that early SARS-CoV-2 introduction contributed to high COVID-19 mortality in Peru. Emphasizing the role of diagnostic confirmation, our study highlights the importance of early detection and accurate testing in managing infectious disease outbreaks.

**Importance:** Latin America was hard hit by the COVID-19 pandemic. Reasons include inadequate healthcare preparation and socio-economic vulnerabilities, likely exacerbated by early undetected SARS-CoV-2 circulation. Diagnostic testing for early SARS-CoV-2 circulation requires exhaustive diagnostic validation due to unspecific reactivity. We used a cohort of circa 1400 febrile patients from August 2019 until May 2021, months earlier than the first seroprevalence study in Lima, Peru, using a two-step diagnostic algorithm. Early 2020 positive samples were further tested with neutralization tests and avidity testing. We confirmed SARS-CoV-2-specific antibodies from April 2020 onwards, suggesting undetected viral circulation circa March 2020, consistent with the first SARS-CoV-2-detection. Early circulation was further confirmed by the significant increase in avidity in positive samples during early 2020 and the modeled peak of reproduction number of >1 during February-March 2020. Using exhaustive diagnostic validation, we detected early SARS-CoV-2 circulation that likely contributed to the severe impact of COVID-19 in Peru.

---

The first cases of COVID-19 were detected in China in mid-December 2019, followed by detections elsewhere in Asia and North America during January 2020 and in Europe during February 2020 (1). The first COVID-19 case in Latin America was detected in Brazil on February 26th, 2020 (1). Despite hosting only 8.5% of the world’s population, Latin America accounted for 30% of COVID-19 deaths worldwide (2). Within Latin America, Peru had the highest per-capita mortality rate both regionally and globally (1, 3). Tentative explanations for the high Peruvian mortality rate have included poorly prepared healthcare systems, genetic predisposition, comorbidities, social inequality and poverty, difficulties following quarantine measures, frequent informal labour, and lack of surveillance systems leading to inaccurate mortality estimates (4). Early SARS-CoV-2 circulation might be a contributing factor to high mortality in Peru.

We investigated SARS-CoV-2-specific IgG antibodies in 1,441 individuals with acute febrile illness investigated between August 2019-May 2021 in the capital city of Lima (**Figure 1A**) by the Peruvian reference laboratory, allowing retrospective analyses several months prior to the first seroprevalence study available from Lima conducted April 2020 (5). The subpopulation consisted of 49.8% females and 50.1% males, with a mean age of 33.7 years and a standard deviation of 20.3 years, comparable to census data from Lima (48.6% males and 51.4% females; mean age=26) (https://censo2017.inei.gob.pe/).

**Figure 1.**
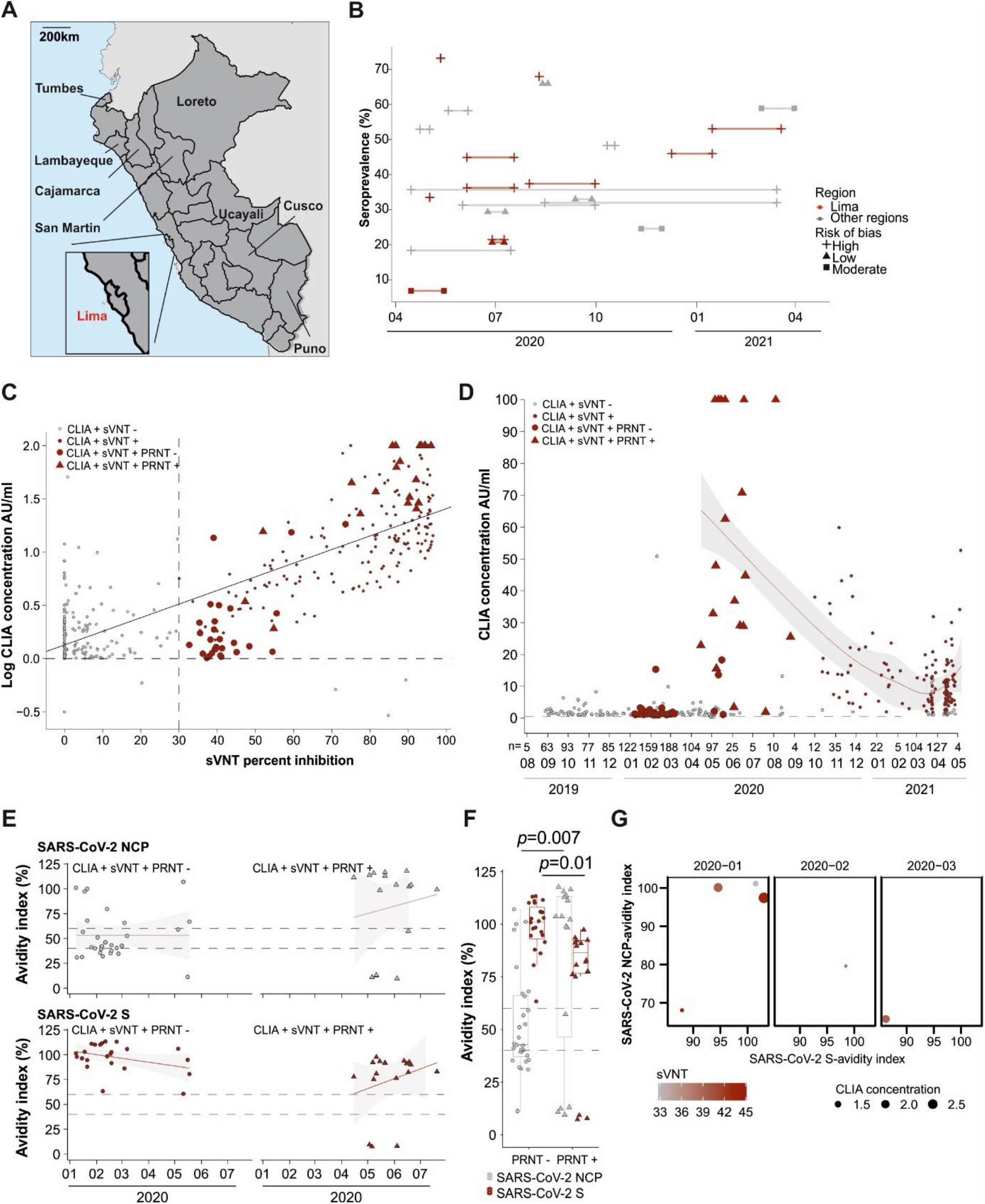
SARS-CoV-2 seroprevalence studies in Peru and diagnostic confirmation. A) Map that shows Peruvian regions where seroprevalence studies have been performed. Data taken from (serotracker.com). B) Seroprevalence studies in Peru, size shows the risk of bias calculated by serotracker, lines connecting the dots show the total study time. C) Reactivity of serum samples to SARS-CoV-2 in a SARS-CoV-2 surrogate virus neutralization test (sVNT), shown in the x axis, and a chemiluminescence immunoassay (CLIA), shown in the y axis. AU/ml, absorbance unit per milliliter. Triangles show early samples that were positive in a plaque reduction neutralization test (PRNT), Big circles show samples that were negative in PRNT. D) CLIA concentration across time in the tested samples. The red line denotes the mean CLIA concentration per month. Gray lines denote the 95% confidence interval. Triangles show early samples that were positive in a plaque reduction neutralization test (PRNT). Big circles show samples that were negative in PRNT. AU/ml: absorbance units per millilitre. E) IgG antibody avidity index against SARS-CoV-2 S and NCP antibodies overtime in PRNT-positive and negative early 2020 samples. Horizontal line denotes the index thresholds of >60% (positive) and <40% (negative) results between these values are considered borderline, as defined by the manufacturer (https://www.euroimmun.com). Gray area denotes the 95% confidence interval. F) Boxplots showing the median avidity by a bar, quartiles by boxes, and minimum and maximum by whiskers. Above, p values of a t-test. G) Reactivity of six serum samples that were SARS-CoV-2 PRNT-negative but SARS-CoV-2 sVNT and CLIA positive, defined as >30 and >1 respectively, as per manufacturer’s instructions. IgG antibody avidity index against SARS-CoV-2 S and NCP antibodies overtime shown in the figure axis, index thresholds of >60% are considered positive as per manufacturer’s instructions.

Approximately 50% of Peruvian seroprevalence studies have a high risk of bias due to the use of potentially non-specific diagnostic tests affected by co-circulating pathogens such as dengue virus (DENV) and *Plasmodium* (**Figure 1B**) (5-7). For confirmation, we used a 2-step testing algorithm including a chemiluminescence immunoassay (CLIA) (SARS-CoV-2 S-RBD IgG kit; Snibe Diagnostic, China), and confirmatory SARS-CoV-2 surrogate virus neutralization test (sVNT; GenScript, USA), both based on the partial SARS-CoV-2 receptor binding domain. Only samples positive in both assays were considered for further analyses to maximize specificity. Additionally, all positive sera from the first six months of 2020, a confirmatory plaque reduction neutralization test (PRNT) based on a 50% reduction in plaque count was performed at 1/8 serum dilution to allow detection of potentially low antibody titers (6). To differentiate between recent and pre-existing SARS-CoV-2 infections, the antibody-antigen binding affinity of SARS-CoV-2 IgG antibodies was determined by conducting an avidity assay using SARS-CoV-2 spike (S1)- and nucleoprotein (NCP)-based IgG ELISAs (Euroimmun, Germany) with an additional 2M urea washing step between sample incubation and detection in one of the replicates.

Several seropositive individuals (n=24) in both CLIA and sVNT were found as early as January-March 2020 (**Figure 1C** and **1D**). These samples showed significantly lower CLIA and sVNT reactivity in comparison to samples from April 2020 onward (CLIA t-test=-6.4, p<0.001; sVNT t-test=-10.9, p<0.001) (**Figure 1C** and **1D**). Additionally, these samples showed a statistically higher mean S-based avidity of 126.4% and lower NCP-based avidity of 11.0% compared to samples from April 2020 onward (S t-test=2.4, p=0.01; NCP t-test=-2.9, p=0.007; **Figure 1E** and **1F**). High avidity to the S protein is reached 1-2 months after SARS-CoV-2 infection and is maintained for approximately 6-8 months (8, 9). High S-based avidity would thus suggest very early SARS-CoV-2 infection in patients sampled during January 2020, contrasting with calculated SARS-CoV-2 phylogenetic introduction estimates and the first SARS-CoV-2 molecular detections in Latin America dating from February 2020 onwards (10). Since PRNT is the serological gold-standard, we further tested all SARS-CoV-2 seropositive samples from January-July 2020 (n=46) using PRNT. None of the early samples from January-March 2020 were PRNT-confirmed, in contrast to 81.8% (18/22) samples from April onwards (Fischer-exact test, p<0.0001). However, a study that tested thousands of convalescent plasma samples using PRNT found that nearly one-third of confirmed SARS-CoV-2 patients that resolved infection had little to no neutralizing activity (11), regardless of when the samples were taken after their COVID-19 infection. Therefore, we cannot exclude that the PRNT-negative (n=5) CLIA, sVNT and S-NCP-high avidity patients in January-March 2020 might have had a past SARS-CoV-2 infection (**Figure 1G**). Of note, no sera that were CLIA positive in 2019 were sVNT positive, suggesting that a minimum 2-step testing algorithm is needed for accurate SARS-CoV-2 serological diagnostics. As a conservative approach, we decided to exclude non-PRNT confirmed samples from January-March 2020 from subsequent analyses, because unspecific reactivity in CLIA and sVNT in those samples seemed the most plausible explanation.

Following diagnostic validation, the SARS-CoV-2 antibody detection rate was 0.9% during April 2020 (1/104; 95% confidence interval (CI), 0.1-5.8%). SARS-CoV-2 antibody detection steadily increased reaching 9.7% (9/93; 95% CI: 5.0-17.8%) in May 2020, reaching a mean of 35% (148/460; 95% CI: 28.0-36.6%) from June afterwards. Given that the mean detection time of SARS-CoV-2-specific IgG antibodies post-symptom onset is 20 days (13), a projected time of infection for the first PRNT-confirmed case could have occurred in early-middle March 2020 which is consistent with the first molecular detection of SARS-CoV-2 in Peru on March 6^th^ (12), 2020 and genomic reconstructions of SARS-CoV-2 in Latin America (10). Avidity to the SARS-CoV-2 NCP and S protein in all PRNT-confirmed CLIA- and sVNT-positive samples from April to July 2020 was high and increased from a mean 77 to 94% for NCP and from 62 to 81% for the S protein by June/July 2020 (NCP t-test=-0.66; p= 0.53; S t-test=-1.12; p= 0.29; **Figure 1E** and **1F**). Those avidity results were thus consistent with onset of SARS-CoV-2 circulation during late February/March 2020.

Onset of SARS-CoV-2 circulation during February/March in Peru was also in concordance with the increase in reported excess mortality peaking mid-March 2020 (**Figure 2A** and **2B**). Stagnation of antibody detection rate during mid-2020 in our study was likely due to the strict non-pharmaceutical interventions (NPIs) in place since March 2020 and mounted population immunity after a high infection peak (**Figure 2B**) (14). Limited transmission is supported by a gradual decline in CLIA titers due to gradual decay of SARS-CoV-2 antibodies (15) during 2020 followed by a resurgence in early 2021 (**Figure 1D**), potentially due to the emergence of antigenic variants such as gamma during early 2021 (1). Notably, resurgence of CLIA titers paralleled an increase in excess mortality and reported COVID-19 cases during early 2021 (**Figure 2A**).

**Figure 2.**
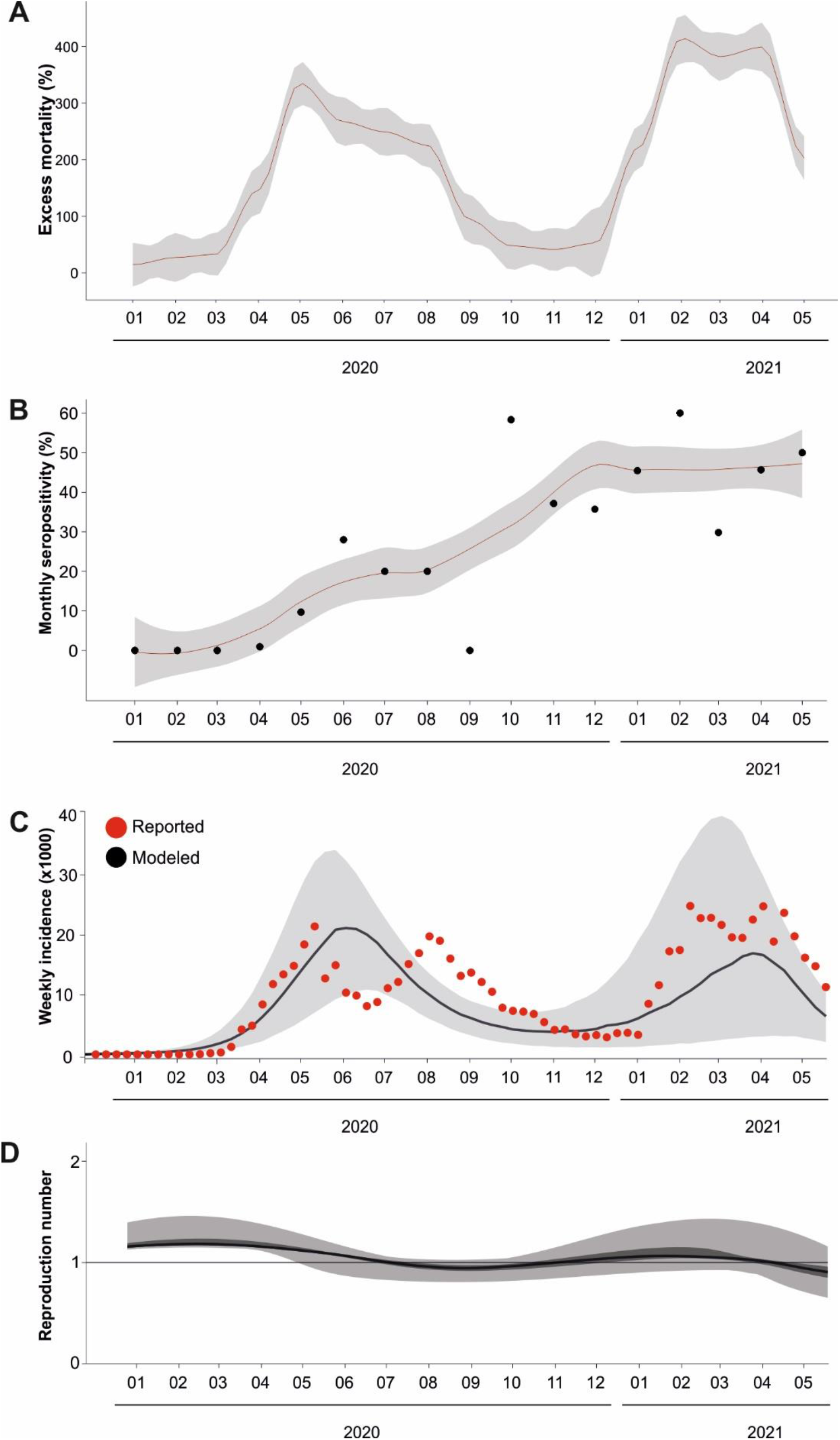
Transmission dinamics of SARS-CoV-2 in Lima, Peru. A) Reported excess mortality. The line shows the monthly reported excess mortality during the study period. Mortality data taken from: https://www.datosabiertos.gob.pe/dataset/fallecidos-por-covid-19-ministerio-de-salud-minsa-. B) Monthly seropositivity from Lima from this study (dots), gray area represents the 95% confidence interval. Linear regression line is shown in red. C) SEIR model showing the reported incidence data (red dots) and the modeled incidence (gray area) calculated using the reported incidence data and antibody detection rate from this study. Weekly incidence data taken from (https://www.datosabiertos.gob.pe/dataset/casos-positivos-por-covid-19-ministerio-de-salud-minsa). The black line denotes the median modeled incidence, and the gray area denotes the 1^st^ and 3^rd^ quartiles of the modeled incidence. D) Effective reproductive number calculated using the model.

Early viral circulation, stagnation, and resurgence of SARS-CoV-2 infections during early 2021 was confirmed using a Susceptible, Exposed, Infected and Recovered Bayesian model relying on reported cases and antibody detection data from our study (16). The model suggested peak virus circulation, with an effective reproduction number (Reff) >1, during February-March 2020, one to two months before the reported peak incidence and mortality which is plausible given the delay in onset of diagnostics estimated at up to 5 days even in affluent settings (16). The model also reconstructed the decrease in virus circulation after July 2020 and increase during end of 2020 (**Figure 2C** and **2D**).

Our data suggest undetected onset of SARS-CoV-2 circulation in Lima during February-March 2020 homologous to other hard-hit Latin American COVID-19 hot spots; e.g., Brazil and Ecuador (1, 2). Considering the intense connectivity between Europe, USA and Peru, it is likely that SARS-CoV-2 circulated before the first case was officially reported.

Limitations of our study include the analysis of a cohort that is not representative of the general population. However, the unique cohort allowed to retrospectively trace SARS-CoV-2 over time during the onset of the pandemic. Another limitation is analysis of sera from febrile individuals whose acute disease may affect serologic diagnostics of SARS-CoV-2, including Malaria and dengue (6, 7). However, our exhaustive serologic analyses allowed robust detection of SARS-CoV-2-specific antibodies.

Our study suggests that early introduction and substantial circulation likely exacerbated the high COVID-19 associated mortality observed in Peru, irrespective of other effect modifiers. Our work emphasizes the complex SARS-CoV-2 serologic diagnostic confirmation in tropical settings, crucial in understanding the pandemic’s trajectory, and highlights the importance of early detection and accurate testing in managing infectious disease outbreaks.

## Data Availability

The code to reproduce the analyses is found in the online repository

## Acknowledgments

The code to reproduce the analyses is found in the online repository: XXXXXXXXXXXX. We thank José Encinas Marroquin, Sebastian Brünink, Arne Kühne and Antje Kamprad. This work was funded by the German federal ministry for economic cooperation and development (BMZ) via the Deutsche Gesellschaft für Internationale Zusammenarbeit (GIZ) GmbH (project number 81262528). The procedures were carried out with the approval of the institutional bioethics committee under the ethic protocol number 6528 from VIA LIBRE and EA2/031/22 from Charité-Universitaetsmedizin Berlin.

## Notes

### Competing Interest Statement

The authors have declared no competing interest.

### Author Declarations

The procedures were carried out with the approval of the institutional bioethics committee under the ethic protocol number 6528 from VIA LIBRE and EA2/031/22 from Charité-Universitaetsmedizin Berlin.

